# Estimates of Actual and Potential Lives Saved in the United States from the use of COVID-19 Convalescent Plasma

**DOI:** 10.1101/2024.05.16.24307505

**Authors:** Quigly Dragotakes, Patrick W. Johnson, Matthew R. Buras, Rickey E. Carter, Michael J. Joyner, Evan Bloch, Kelly A. Gebo, Daniel F. Hanley, Jeffrey P. Henderson, Liise-anne Pirofski, Shmuel Shoham, Jonathon W. Senefeld, Aaron AR Tobian, Chad C. Wiggins, R. Scott Wright, Nigel S. Paneth, David J. Sullivan, Arturo Casadevall

## Abstract

In the Spring of 2020, the United States of America (USA) deployed COVID-19 convalescent plasma (CCP) to treat hospitalized patients. Over 500,000 patients were treated with CCP during the first year of the pandemic. In this study, estimated the number of actual inpatient lives saved by CCP treatment in the USA based upon CCP weekly use, weekly national mortality data, and CCP mortality reduction data from meta-analyses of randomized controlled trials and real-world data. We also estimate the potential number of lives saved if CCP had been deployed for 100% of hospitalized patients or used in 15% to 75% of outpatients. Depending on the assumptions modeled in stratified analyses, CCP was estimated to have saved between 16,476 and 66,296 lives. The CCP ideal use might have saved as many as 234.869 lives while preventing 1,136,133 hospitalizations. CCP deployment was a successful strategy for ameliorating the impact of the COVID-19 pandemic in the USA. This experience has important implications for convalescent plasma used in future infectious disease emergencies.

**Significance statement:** When the COVID-19 pandemic struck in 2020, the population lacked immunity, no validated therapies were available, and mortality was high. COVID-19 convalescent plasma (CCP) was authorized in the United States for treatment of hospitalized patients based on historical evidence of convalescent plasma (CP) efficacy and findings from a nationwide registry suggesting that it reduced mortality. However, this decision was controversial because it was not based on evidence from randomized controlled clinical trials. In this study, we leveraged CCP use and mortality data combined with CCP efficacy data to show that CCP reduced mortality and saved tens of thousands of lives the first year of the pandemic. This provides a powerful basis to consider CP deployment in future infectious disease emergencies.

## Introduction

In the spring of 2020, the United States of America (USA) faced a rapidly worsening coronavirus disease 2019 (COVID-19) pandemic caused by a novel infectious agent, SARS-CoV-2. In the absence of specific therapies for COVID-19, the USA Food and Drug Administration made COVID-19 convalescent plasma (CCP) available in 2020, first under compassionate use in late March, then under an Expanded Access Program (EAP) and registry in early April, and finally under Emergency Use Authorization (EUA) in late August (1). CCP was qualified initially based on the donors having had a previously positive SARS-CoV-2 nucleic acid test, not on specific antibody levels. The EAP registry enrolled approximately 105,000 patients by late August 2020 (1) and produced early evidence of safety (2, 3) and efficacy (4, 5). By the Fall of 2020 as many as 40% of hospitalized patients were being treated with CCP (6). However, disappointing results from several randomized controlled trials (RCTs) assessing CCP efficacy in hospitalized patients in India (7), Argentina (8), the United Kingdom (9), and Italy (10), combined with the availability of small molecule antivirals in the form of remdesivir, led to a substantial decline in use by early 2021; we previously estimated this decline was associated with as many as 30,000 excess deaths by mid-2021 (6).

In retrospect, early RCTs examining CCP efficacy in hospitalized patients were unlikely to show benefit because of design flaws that included use of plasma with inadequate specific antibody concentrations, inexact endpoints, late CCP administration (*e*.*g*. use during the inflammatory phase rather than the viral phase of COVID-19), and/or insufficient power (11, 12). The early phase of the pandemic in the USA also precluded a number of factors vital for conduct of successful RCTs including: (i) training of sites and site initiation visits; (ii) precise pre-deployment of CCP; (iii) a moving pandemic that affected different geographic regions differently; (iv) impaired access to research staff due to work lockdowns ; and (v) the lack of a national network to conduct pandemic related research smoothly and seamlessly. Although not known at the time, later retrospective analysis of EAP data showed that distance between CCP collection and use also reduced efficacy (13), adding another variable that could have influenced the outcome of some RCTs. Subsequent trials of CCP using units with high levels of spike-protein specific IgG (high titer CCP) early in disease eventually established its efficacy (14, 15). However, by the time this information was available, rapid acquisition of antibody immunity from natural infection and vaccination in the general population, combined with widespread availability of small molecule antiviral agents and monoclonal antibodies (mAbs), lowered the demand for this passive antibody therapy. Nonetheless, CCP has retained a role in the COVID-19 therapeutic armamentarium in immunosuppressed patients, in whom, even in the first year of the pandemic there was evidence for efficacy (16). With the loss of mAb efficacy due to continued SARS-CoV-2 evolution (17), CCP is again the only available antibody-based with activity against SARS-CoV-2 (18).

Four lines of evidence show that CCP reduces COVID-19 inpatient mortality when used early in disease: 1) registry data from the USA (5), Argentina (19) and Italy (20); 2) real world data from use in the USA (21); 3) a meta-analysis of over 30 RCTs (22); and 4) epidemiologic data showing a strong negative correlation between CCP use and mortality, with a reciprocal relationship between weekly use and the national death rate (6). From the available epidemiological data, it was estimated that had the USA not deployed CCP in 2020, approximately 96,000 additional deaths would have occurred during the first year of the pandemic (6). In the present analysis we revisit the question of how CCP use affected overall USA mortality by combining CCP usage data with mortality statistics and efficacy measures from RCTs and real-world data.

## Results

### Actual lives saved in hospitalized patients

Although most patients hospitalized with COVID-19 had progressed past the interval of optimal CCP efficacy, virtually all CCP used in the USA involved hospitalized patients, reflecting the initial EUA restriction to inpatient use. Only in December 2021, after an outpatient RCT revealed efficacy (14), did the FDA authorize outpatient use, and then only in immunosuppressed patients. Using the 647,795 CCP units dispensed from July 2020 to March 2021 as a measure of the number of patients treated and applying the mortality reduction measures from various published studies (21, 22), we calculated that CCP deployment in the United States saved between 16,585 to 67,706 lives in this period of the pandemic (Table 1). Importantly, even with the conservative estimate of mortality reduction of 13%, a significant number of lives were saved (95% Credible Interval (CI): 4,356 to 36,032) (Table 1). The range in crude estimates reflects the different assumptions and methods used in calculating the estimate. Although this range is large, all models converge upon the conclusion that CCP saved lives, as indicated by the credible intervals.

**Table 1.**
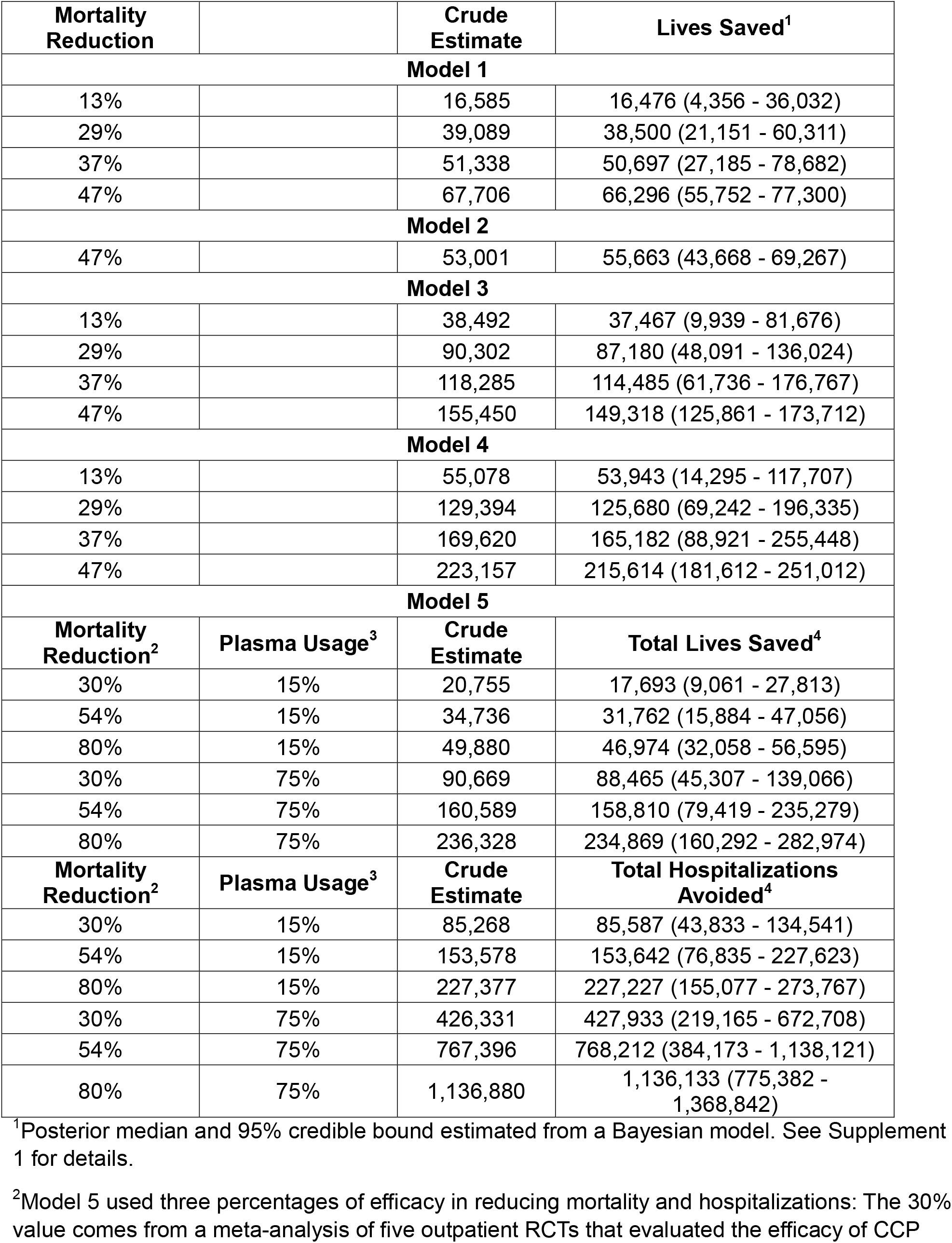
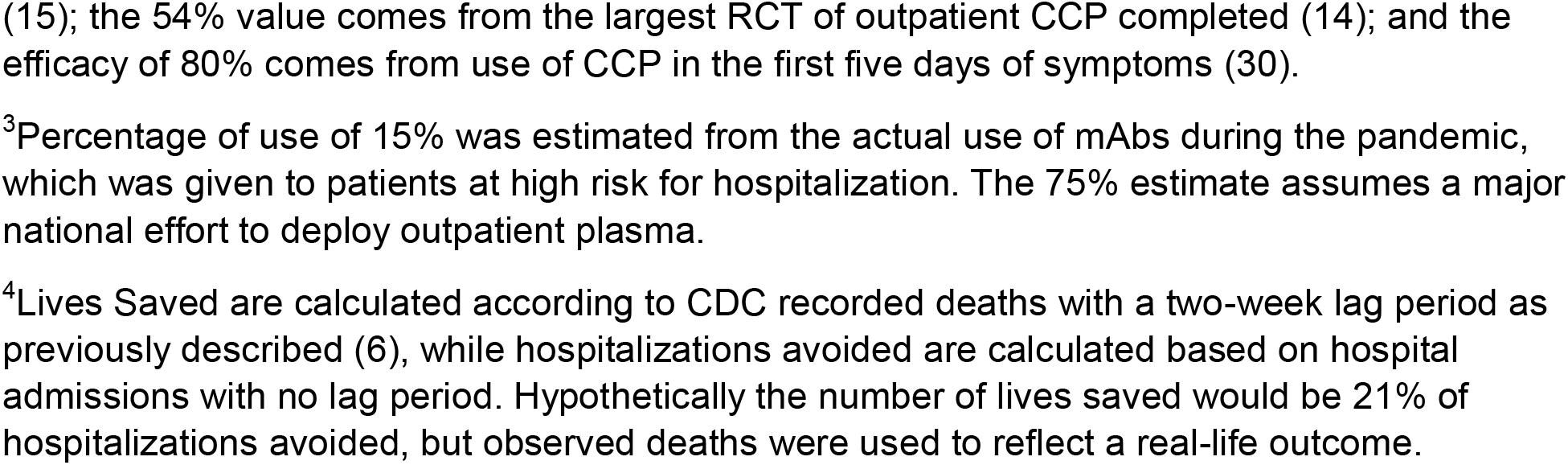
Estimates of lives saved from the deployment of CCP in the USA.

| **Mortality Reduction** |  | **Crude Estimate** | **Lives Saved<sup>1</sup>** |
| --- | --- | --- | --- |
| **Model 1** |  |  |  |
| 13% |  | 16,585 | 16,476 (4,356 - 36,032) |
| 29% |  | 39,089 | 38,500 (21,151 - 60,311) |
| 37% |  | 51,338 | 50,697 (27,185 - 78,682) |
| 47% |  | 67,706 | 66,296 (55,752 - 77,300) |
| **Model 2** |  |  |  |
| 47% |  | 53,001 | 55,663 (43,668 - 69,267) |
| **Model 3** |  |  |  |
| 13% |  | 38,492 | 37,467 (9,939 - 81,676) |
| 29% |  | 90,302 | 87,180 (48,091 - 136,024) |
| 37% |  | 118,285 | 114,485 (61,736 - 176,767) |
| 47% |  | 155,450 | 149,318 (125,861 - 173,712) |
| **Model 4** |  |  |  |
| 13% |  | 55,078 | 53,943 (14,295 - 117,707) |
| 29% |  | 129,394 | 125,680 (69,242 - 196,335) |
| 37% |  | 169,620 | 165,182 (88,921 - 255,448) |
| 47% |  | 223,157 | 215,614 (181,612 - 251,012) |
| **Model 5** |  |  |  |
| **Mortality Reduction<sup>2</sup>** | **Plasma Usage<sup>3</sup>** | **Crude Estimate** | **Total Lives Saved<sup>4</sup>** |
| 30% | 15% | 20,755 | 17,693 (9,061 - 27,813) |
| 54% | 15% | 34,736 | 31,762 (15,884 - 47,056) |
| 80% | 15% | 49,880 | 46,974 (32,058 - 56,595) |
| 30% | 75% | 90,669 | 88,465 (45,307 - 139,066) |
| 54% | 75% | 160,589 | 158,810 (79,419 - 235,279) |
| 80% | 75% | 236,328 | 234,869 (160,292 - 282,974) |
| **Mortality Reduction<sup>2</sup>** | **Plasma Usage<sup>3</sup>** | **Crude Estimate** | **Total Hospitalizations Avoided<sup>4</sup>** |
| 30% | 15% | 85,268 | 85,587 (43,833 - 134,541) |
| 54% | 15% | 153,578 | 153,642 (76,835 - 227,623) |
| 80% | 15% | 227,377 | 227,227 (155,077 - 273,767) |
| 30% | 75% | 426,331 | 427,933 (219,165 - 672,708) |
| 54% | 75% | 767,396 | 768,212 (384,173 - 1,138,121) |
| 80% | 75% | 1,136,880 | 1,136,133 (775,382 - 1,368,842) |

<sup>1</sup>Posterior median and 95% credible bound estimated from a Bayesian model. See Supplement 1 for details.
<sup>2</sup>Model 5 used three percentages of efficacy in reducing mortality and hospitalizations: The 30% value comes from a meta-analysis of five outpatient RCTs that evaluated the efficacy of CCP
(15); the 54% value comes from the largest RCT of outpatient CCP completed (14); and the efficacy of 80% comes from use of CCP in the first five days of symptoms (30).
<sup>3</sup>Percentage of use of 15% was estimated from the actual use of mAbs during the pandemic, which was given to patients at high risk for hospitalization. The 75% estimate assumes a major national effort to deploy outpatient plasma.
<sup>4</sup>Lives Saved are calculated according to CDC recorded deaths with a two-week lag period as previously described (6), while hospitalizations avoided are calculated based on hospital admissions with no lag period. Hypothetically the number of lives saved would be 21% of hospitalizations avoided, but observed deaths were used to reflect a real-life outcome.

### Potential lives saved with optimal CCP deployment

We next estimated the hypothetical efficacy of CCP treatment if infrastructure had already been in place to collect, manufacture, and distribute high titer CCP to 100% of hospitalized patients within 3 days of admission. Depending on the COVID-19 mortality estimate Models 3 and 4 yield from 37,467 to 149,318 and 53,943 to 215,614 lives saved, respectively, each of which would be statistically significant based on the credible intervals (Table 1).

Using data from five outpatient RCTs (15), it is possible to estimate the effect of CCP on mortality had this therapy been authorized for outpatient use in the early days of the pandemic. However, outpatient deployment would have required specialized infrastructure that was not immediately available at the time. Furthermore, some physicians were concerned about potential side effects such as antibody-dependent enhancement and antibody-triggered cytokine storms (23). Early outpatient use of CCP would have required a monitored environment similar in some ways to the inpatient environment (24). But by May 2020 (2), we had learned that CCP is a safe inpatient therapy (25), and by Fall 2020 it had been used successfully in an outpatient RCT (26) without safety concerns (27).

Although the logistics of outpatient CCP use are more complicated than in-hospital use (24), successful deployment of outpatient mAb therapy and the availability of outpatient RCT data (14, 28) established the feasibility of this option in the USA. Efficacy of outpatient use of CCP was estimated in three ways: a 30% reduction in hospitalization based on a meta-analysis of five trials (29); a 54% reduction based on findings of the largest RCT (14); and an 80% reduction based on findings from the subset treated within 5 days in the largest RCT (30). However, the more complex logistics of outpatient CCP use make it unlikely that everyone at risk for progression would have received this therapy as only 15% of eligible patients received mAb outpatient therapy (31). Had a similar percentage of high-risk individuals been treated with CCP in the first year of the pandemic, we estimate that between 85,268 and 227,377 hospitalizations could have been avoided, depending on the efficacy estimate. Using the 21% overall mortality rate for hospitalized patients at that time, this would have further prevented about 17,693 to 46,974 deaths, depending on the efficacy estimate, since most deaths from COVID-19 occurred in hospitals (Figure 1, Table 1).

**Figure 1.**
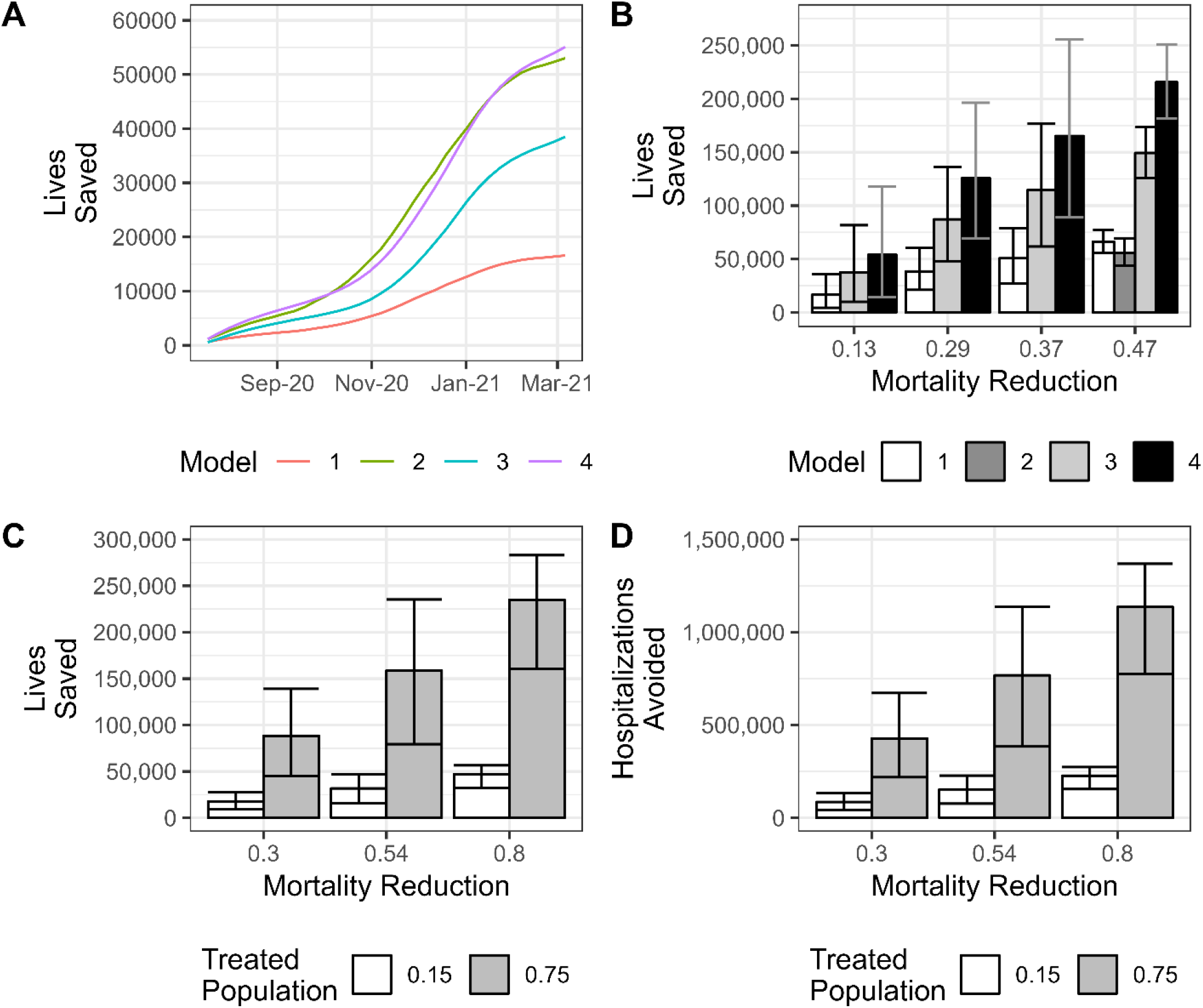
Bayesian estimates of total lives saved with confidence intervals given various models of CCP usage and efficiency from July 2020 through March 2021. **A**. Summations of estimated lives saved using the most conservative parameters of each model as a function of time throughout the entire period. **B**. Estimated lives saved in models 1 through 4 with confidence intervals depicted by error bars. **C**. Estimated lives saved in model 5 with confidence intervals depicted by error bars. **D**. Estimated hospitalizations avoided in model 5 with confidence intervals depicted by error bars.

Reduction in hospitalizations would have also reduced stress on the health care system, which itself was associated with 2,000-80,000 additional deaths from causes other than COVID-19 in the first year of the pandemic (32). These estimates suggest that the secondary effects in reducing hospital stress might have saved additional lives, increasing our estimates of lives saved according to Model 1 (Table 1) from a minimum of 18,476 (16,476 + 2000) to a maximum of 146,296 (66,296 + 80,000). Had public health and medical authorities been able to provide CCP to 75% of high-risk patients (Model 4), these numbers would have risen to between 55,943 (53,943 + 2000) to 395,6147 (215,614 + 80,000). With 407,100 USA deaths during the first year of the pandemic, such a deployment would have reduced mortality by 13-72% and substantially mitigated the impact of the pandemic in the USA. Given an average hospitalization cost of $41,000 per patient (33) and an average cost of $750 per unit of CCP, we estimate outpatient deployment with treatment of only 15% of eligible patients, with a 54% reduction in progression to hospitalization (14), would have saved the USA approximately $6 billion. If given to 75% of eligible patients, savings would approach $31 billion.

Figure 1 shows estimated lives saved with different mortality reduction assumptions and potential lives saved had universal CCP use been instituted for hospitalized patients. Because it is uncertain which mortality reduction value is most tenable, we opted to present all the estimates in Table 1 and the most conservatives estimates only in Figure 1. Despite these variations, all estimates show that thousands of lives were saved by CCP deployment.

### Safety of CCP

Intrinsically linked to the conclusion that CCP saved lives is the assumption that transfusion of CCP is safe. Numerous observational studies and RCTs have established that CCP is a safe therapy (34). However, like all generally safe drugs such as penicillin that can occasionally trigger fatal reactions (35), plasma administration was associated with severe reactions on rare occasions. The standard transfusion reactions - transfusion related acute lung injury (TRALI) and transfusion associated circulatory overload (TACO) were very rare, and while antibody-dependent enhancement was feared, it was not observed (2, 3). TRALI occurs after transfusion in 1 of 2000 plasma components and is fatal in 5-10% of cases (36-38). Among 20,000 individuals who received CCP there were 36 reports of TACO, 21 reports of TRALI and 21 reports of severe allergic transfusion reactions, which was similar to complication rates associated with infusion of fresh frozen plasma (3), of which about 2,000,000 units are transfused in the USA each year primarily to provide replacement of coagulation factors (39). At least one fatal reaction to CCP infusion has been described in the literature (40). When considering presumptive severe reactions from CCP administration occurring in critically ill patients, it is often difficult to distinguish these from worsening of the underlying illness, especially in the face of concurrent pneumonia, ARDS, ongoing mechanical ventilation, ventricular dysfunction, and arrhythmias. Nevertheless, in our estimates we sought to consider the worst possible scenario for CCP in contributing to COVID-19 related deaths to provide the most conservative estimate of lives saved. The EAP registry recorded 63 deaths among 20,000 individuals transfused with CCP within 4 h of plasma infusion, of which 10 were judged as possibly related to CCP. Extrapolating this mortality rate to our study, given that 647,795 units were administered, would mean that 32 to 324 deaths from CCP would have to be subtracted from the total number of lives saved.

### A model for how CCP reduced mortality in COVID-19

A causal association between CCP usage and lives saved is strengthened by an understanding of the CCP mechanism of action. CCP administration has been shown to reduce SARS-CoV-2 viral load in macaques (41), hamsters (42, 43), and mice made susceptible to this coronavirus by expressing the human angiotensin-converting enzyme 2 (ACE2) receptor (44, 45). In hospitalized patients, administration of CCP with greater neutralizing antibody content was associated with greater SARS-CoV-2 viral load reduction (46). Both animal and clinical studies thus establish CCP as an antiviral therapy, consistent with the accepted view that specific antibody can neutralize viral particles in vivo. For both CCP and mAb preparations, the active ingredient against SARS-CoV-2 is a specific antibody. Consistent with the antiviral activity of both preparations, monoclonal antibody RCTs reported increased rates of viral clearance in the intervention arms (47), confirming the efficacy of specific antibody as an antiviral agent.

Dose response effects are powerful tools for establishing causality in science and medicine (48). In this regard, several studies reported dose-response effects between CCP specific antibody content and favorable clinical outcomes (5, 19, 29, 49-52). Greater viral load reduction was also observed in hospitalized patients receiving greater quantities of CCP (two units) in an RCT (53). Given that specific antibody is an effective antiviral, greater efficacy for CCP units with higher specific antibody content can be expected to mediate stronger antiviral effects, that should translate into favorable outcomes.

A third line of evidence for a causal association between CCP use and reduced mortality comes from its effects on inflammatory markers, which served as surrogate markers of COVID-19 severity. CCP administration was associated with a reduction in markers of inflammation, including C-reactive protein (54-56) and IL-6 (54, 57-59). Since increased levels of IL-6 correlate with increased mortality and anti-IL-6 therapy reduces COVID-19 mortality (60), CCP-associated reductions in IL-6 could have contributed to its effect on mortality. The anti-inflammatory effect of CCP could be a consequence of its antibody-mediated antiviral effect where reduced viral load elicits less inflammation and/or other components (61). Most patients with COVID-19 die because of profuse pulmonary inflammation that impairs gas-exchange (62). In a Belgian RCT, CCP transfused within 48 hours of mechanical ventilation reduced deaths(63). Consequently, CCP anti-inflammatory effects can be incorporated into a model for mortality reduction whereby reduced CCP reduces viral load and inflammatory cytokines and thus lowers the probability of disease progression to end stage pulmonary compromise (Figure 2). In this regard, viral clearance from both small molecule antivirals and specific antibody is a surrogate for clinical efficacy in preventing progression of disease (47). Consistent with the critical role of specific antibody in host defense, the absence of antibody to SARS-CoV-2 is a poor prognostic marker associated with increased mortality in COVID-19 (64, 65), which provides an additional explanation how the administration of CCP reduced mortality by providing recipients with antibody to the virus.

**Figure 2.**
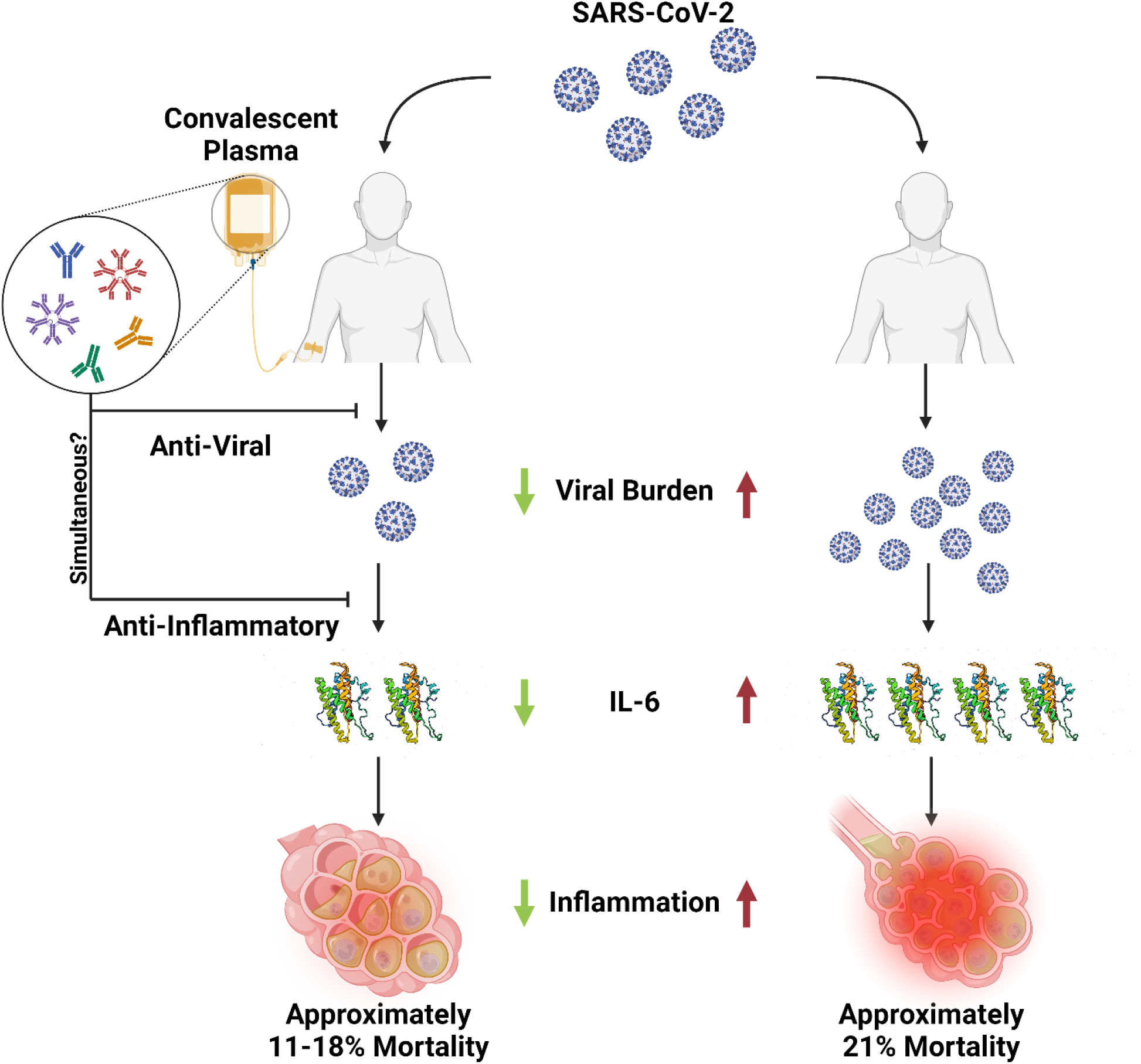
Proposed scheme for the reduction of COVID-19 mortality by CCP. In the USA CCP was used almost exclusively in hospitalized patients, of whom the majority were admitted because of some pulmonary compromise. Hence, the reduced mortality described here is proposed to reflect the subset that were sufficiently early in the course of disease such that the administration of antibody could modify the progression of disease to result in better outcomes. CCP has been shown to have antiviral activity and to be associated with reduced inflammatory mediators including IL-6. According to this scheme, CCP administration led to reduced inflammation that translated into lower mortality for a subset of treated hospitalized patients. Created with BioRender.com

## Discussion

Our estimates indicate that CCP deployment in the USA in 2020 saved thousands of lives. This public health benefit is in alignment with the decisions to authorize its use during a national emergency, particularly early in the pandemic when there was a great need for effective therapies. Furthermore, these data support the use of this therapy in future infectious disease outbreaks. Our results suggest that, had more CCP use been encouraged, and had its availability been prioritized by medical and governmental authorities, more lives would have been saved. Despite receiving emergency use authorization by the FDA in August 2020, CCP use was not often recommended by guideline committees for COVID management, which held out for RCT data before making recommendations, but such evidence was not available early in the pandemic. Had CCP been universally deployed in hospitals, as was done for supplemental oxygen and corticosteroids in hypoxic individuals, we estimate that the total lives saved among hospitalized patients would have increased ranging from 36,838 to 215,195 depending on the model used and the assumed efficacy. Universal use would not have been possible in the early days of the pandemic when CCP was scarce but by the Fall of 2020 supplies were plentiful and up to 40% of hospitalized patients in the USA were receiving CCP (6). COVID-19 was particularly devastating for residents of long-term care facilities (66). Mortality rates due to COVID-19 in these facilities were particularly high (67) and CCP deployment may have had an outsized impact upon this population.

In considering our estimates, we acknowledge several limitations of the analysis. The number of CCP units used for the calculations provided by the BCA does not capture all the CCP used in the USA, particularly in the early days of the pandemic when some CCP was sourced locally. While the exact number of units used is unknown, the estimates used in this study capture the great majority of CCP used in the USA. The mortality reduction estimates used to calculate the lives saved varied widely and the extent to which they resembled use and efficacy in more 2,000 clinical settings that used CCP throughout the USA is uncertain. Of note, CCP efficacy was found to vary with distance between donor collection to patient administration sites, with a significant reduction in efficacy when the distance exceeded 150 miles, likely reflecting donor-recipient mismatches arising from local viral evolution (68), a phenomenon consistent with geographic antigenic variation by SARS-CoV-2 (13). We did not model this distance effect on the potential of CCP for saving lives. Had all CCP been locally sourced our estimates of lives saved would have been higher.

In a previous epidemiologic study using regression analysis of USA population data correlating weekly mortality figures with CCP use, CCP deployment was estimated to have saved about 96,000 lives in the first year of the pandemic (6). The difference in lives saved between the epidemiologic study and the modeling estimates of the present study could arise from lower efficacy in hospitalized populations studied in RCTs or from trial-associated methodological differences in CCP administration. For example, RCTs inevitably included enrollment and randomization protocols that may have further delayed the administration of CCP, thus reducing its efficacy (11). Additionally, epidemiological analyses could have overestimated the lives saved if the assumptions used to correlate overall mortality with CCP usage did not account for possible confounders. Finally, this previous model estimated a base mortality of 25% which is notably higher than the approximately 21% estimated in this study and the referenced literature, and the higher the base mortality the more lives saved. Nevertheless, both the prior (6) and current analyses are consistent in concluding that CCP deployment saved tens of thousands of lives.

Despite the apparent success of CCP in lowering COVID-19 mortality in the USA, we note that many aspects of its deployment were suboptimal. Early in the pandemic the ability to test donated plasma for antibody content was limited and many patients received units with little or no specific antibody to SARS-CoV-2 (69, 70). In a future emergency where public health authorities are again confronted with a situation where it is difficult to ascertain antibody levels they might consider using two plasma units from separate donors to increase the probability of providing sufficient specific antibody to the recipient (71). Once antibody levels can be determined, the optimal units for plasma therapy should be those in the upper 2-3 deciles of geometric mean antibody levels, which after a ten-to-twenty-fold dilution are still in the protective range (71). In addition, many patients in the first year of the pandemic were treated after three days of hospitalization (72), when CCP administration was likely to have little or no effect on outcome (5). The COVID-19 pandemic has yielded voluminous information on effective use of passive antibody therapies that reinforce the historical evidence (11), including the importance of using them early in the course of disease (29), the efficacy immunocompromised individuals (18), and the need to use units with high pathogen-specific immunoglobulin content (51).

In less than a quarter of this new century, humanity has confronted no fewer than seven major viral outbreaks with pandemic potential: Severe Acute Respiratory Syndrome (SARS) in 2003, Middle Eastern Respiratory Syndrome (MERS) in 2008, Influenza H1N1 in 2009, Ebola virus (2013), Zika virus in 2015, SARS-CoV-2 in 2019 and mPox in 2022. For SARS (73), MERS (74), influenza H1N1 (75), Ebola (76), SARS-CoV-2 (this study) and mPox (77), convalescent plasma (CP) was either used clinically or considered. The USA experience with CCP provides a roadmap for future deployments of convalescent plasma (CP). Our models demonstrate that the use of CP at least as a stopgap measure until additional treatments are developed and mobilized should be considered part of pandemic preparedness. In addition, our estimates provide robust evidence that preparedness for a future pandemic includes an outpatient infrastructure that can facilitate early delivery of high titer CP. As was the case with COVID-19, CP is likely to be the only pathogen-specific therapy available for new infectious disease until drugs, mAbs and vaccines become available. The long record of serotherapy efficacy dates to its first use in the 1890’s for diphtheria management (78) and includes efficacy during the 1918 influenza pandemic (79). The availability of CP as soon there are survivors supports CP use while safety and efficacy data are obtained as was permitted by the EAP in the USA (1).

The careful recording of the results of CP deployment in a registry such as the EAP (1) provides information on this therapy which can inform the design of RCTs if necessary. RCTs of CP efficacy should not be launched until the optimal dose and timing of the intervention is established. Without this information, one runs the of risk of misleading negative trials using suboptimal treatment, as occurred frequently in the early CCP trials (80). The argument that CP deployment inhibits the conduct of RCTs is mistaken; at least five RCTs were completed in the USA while CCP was available as part of the EAP and its subsequent use under the EUA (80). Our analysis provides reassurance that FDA decisions on the deployment of CCP and the enormous efforts made by physicians, blood bankers, and the public in securing plasma in the first year of the pandemic saved thousands of lives.

## Methods

The overarching goal of the analysis was to estimate the lives saved based on the availability of CCP. To achieve this objective, we developed several models based on available CCP use and mortality data from 7/18/20 through 3/6/21. Each of these models was selected to test various assumptions about how the public health benefit could be measured based on published studies examining the efficacy and national trends in hospitalizations and CCP utilization. Crude estimates of the lives saved were obtained through direct computation using the modeling frameworks described below. The specified models and crude estimates were then combined with Bayesian estimation to produce credible bounds to measure the precision in the estimates (see Supplement 1 for details on the Bayesian modeling). The following sections detail how the hyperparameters and modeling frameworks were selected.

### CCP units used and patients treated

The number of CCP units dispensed in the USA in the first year of the pandemic was obtained from the Blood Centers of America Inc (BCA, West Warwick, RI), based on the reported number of units shipped from all blood supplies to hospitals nationwide (6). This number does not capture CCP produced by independent hospitals and transfusion centers (6) as some CCP was collected and processed locally, as previously described (53, 81). Nevertheless, BCA data represents approximately 90% of all units given in the US. Given that the USA FDA recommendations for CCP use in 2020 were to use one unit per patient, our estimates assumed that the number of units used corresponded to the number of patients treated.

### CCP mortality reduction percentages

We made two estimates of this parameter – one based on RCT’s and propensity matched studies, and another based on real world data. From a meta-analysis of all controlled studies through 2022 (39 RCTs with 21,529 participants; 70 propensity matched cohort studies with 50,160 participants) we estimate that CCP reduced mortality by 13% in all hospitalized patients and by 37% in inpatients treated early with high titer units (22).

Using real world data, CCP was estimated to reduce mortality in all hospitalized patients by 29% and by 47% when high titer units were used early in hospitalization (21). These mortality ranges include the most recent RCT in hospitalized patients reporting a 21% reduction in mortality (63), published after the above meta-analysis. Justification for the assumption of early in-hospital use comes from Mozaffari et al (72), who reported that by Fall 2020 over 83% of a large sample of patients in the United States treated with CCP were being treated in the first three days of hospitalization. Confidence intervals reported in these studies were used to generate prior distributions in the Bayesian framework.

### Estimating hospitalized lives saved by deployment of CCP

The weekly number of hospitalized individuals, weekly deaths associated with COVID-19 estimated as previously described (6), and weekly hospital admissions were acquired from the United States Centers for Disease Control and Prevention (CDC) COVID-19 reporting databases. The proportion of early administered CCP was calculated according to Mozaffari et al (72) who provided the percentages of individuals treated by hospital day in a database representing 20% of all USA hospitals.

Lives saved by CCP were calculated using the four separate estimates of mortality benefit conferred by CCP shown above in hospitalized patients (22), i.e. 13% or 29% for any treatment in hospitals; 37% or 47% if treatment was early with high titer plasma (21).

**Model 1**. Evaluates the question: how many lives did CCP save in comparison to a situation where CCP was never used? In this scenario:

Total Deaths = (Untreated Patients ^*^ Untreated Mortality%) + (Treated Patients ^*^ Treated Mortality%)

We estimated the untreated mortality each week by substituting that term with [Recorded Deaths / (Admissions - Treated Patients * Mortality Reduction)]. We then calculated the lives saved as the difference between the above and (Admissions * Untreated Mortality) where the comparison is to the absence of CCP treatment, using the four mortality reduction fractions described above from trial and real-world data (*i*.*e*. 13%, 29%, 37% and 47%) obtained from (21, 22).

**Model 2**. This model for estimating actual lives saved differs from Model 1 in that we added consideration of optimal use of plasma, *i*.*e*. in the first three days of hospitalization. For this estimate we used the timing of CCP administration as reported by Mozzafari (72), who reported that by December 2020, in a sample of 20% of US hospitals, 83% of patients were receiving CCP in the first three days of hospitalization. We used the real-world efficacy data from Arnold *et al*. (21) of a 47% reduction in mortality if given in the first 3 days and no efficacy if used thereafter. We assumed that post-December 2020 usage resembled rates observed in December as USA physicians had apparently learned the need to use it early in the course of hospitalization and CCP was plentiful. The lives saved estimated from this model was calculated using the same methodology as model 1 except that the number of treated patients each week and the untreated mortality rate were recalculated according to estimated early plasma use.

**Models 3 and 4** estimate the number of lives that would have been saved had CCP been administered to 100% of hospitalized patients, using the four measures of efficacy in reducing mortality described above. Both models are similar to model 1 except for the assumption that all hospitalized patients received CCP.

Total Deaths = (Admissions) * (Treated Mortality%)

Model 3 Total Lives Saved = Recorded Deaths – (Admissions ^*^ Treated Mortality%)

Model 4 Total Lives Saved = (Admissions * Untreated Mortality) – (Admissions * Treated Mortality%)

Models 3 and 4 both compare the number of deaths that we estimate could have been saved if all hospitalized patients had been treated in the first three days but differ in the way deaths were estimated. Model 3 uses a weighted estimate of 21% average mortality based on a regression analysis of weekly death rates previously established (6). Model 4 uses the actual number of deaths reported by the USA CDC, synchronizing these to the number of admissions with a two-week lag to allow for deaths to occur. These assumptions add different uncertainties. The accuracy of Model 3 is dependent on a regression analysis estimate while in Model 4 not all deaths occurred exactly two weeks after admission and the model does not account for the proportion of patients who did receive CCP, since the USA CDC mortality numbers reflect all who died including those treated with CCP.

### Model 5: Estimating potential lives saved had CCP been deployed for outpatient use

Given the greater efficacy of CCP when used early in the course of disease is likely that outpatient use could have saved even more lives than inpatient use. A RCT of CCP outpatient efficacy early in the pandemic reported a 48% relative risk reduction in progression to severe illness likely to lead to hospitalization in elderly patients (28). Subsequently, a large RCT of CCP outpatient use reported a 54.3% efficacy in reducing hospitalization (14). Consequently, we estimated the potential lives saved by outpatient use based on outpatient CCP efficacy data obtained during the pandemic. When CCP was given in the first five days of symptoms its efficacy in reducing progression to hospitalization rose to 79.9%, similar to monoclonal antibodies (30). A more conservative figure of 30% for outpatient CCP emerges from a meta-analysis of five RCTs including international trials (29). We used all three estimates – 30%, 54% and 80% - as shown in Table 1. Although not all patients who died of COVID-19 died in hospitals, the vast majority did (82). Consequently, it is possible to estimate lives saved by deployment of outpatient CCP since individuals not admitted to hospital were assumed to contribute little to the overall death rate. In this estimate, the number of lives saved is seen as proportional to the number of hospitalizations avoided, assuming that the mortality rate would otherwise be unchanged in the hospitalized proportion of patients:

Total lives saved = Recorded Deaths * proportion of patients treated * efficacy of CCP

## Data Availability

All data produced in the present study are available upon reasonable request to the authors

