## Supplemental File 1 for "Estimates of Actual and Potential Lives Saved in the United States from the use of COVID-19 Convalescent Plasma"

### Supplemental Methods

### 1 Generating Prior Distributions

#### 1.1 Deriving alpha and beta paramaters from RRR/RR

The treatment benefits of COVID convalescent plasma (CCP) are generally reported as either relative risk reduction (RRR) or relative risk (RR) in the form of point estimates and 95% confidence intervals. In this analysis, mortality/hospital admission reduction is modeled using a $Beta(\alpha, \beta)$ distribution, which allows us to incorporate uncertainty in the estimates derived from the models. In cases where RR is reported, RRR is first calculated based on $RRR = 1 - RR$ and is considered the mean estimate. From here, the standard deviation of the distribution can be calculated using the limits from the 95% CI using the following formula:

$$SD = \frac{\text{upper CL} - \text{lower CL}}{2 \times 1.96}, \text{where CL=confidence limit} \tag{1}$$

The variance of this distribution is then determined as:

$$\text{Var} = SD^2 \tag{2}$$

The $Beta$ parameters can then be calculated as follows:

$$\alpha = \left(\frac{1 - \text{Mean}}{\text{Var}} - \frac{1}{\text{Mean}}\right) \times \text{Mean}^2 \tag{3}$$

$$\beta = \alpha \times \left(\frac{1}{\text{Mean}} - 1\right) \tag{4}$$

#### 1.2 ARR to RRR

In cases where absolute risk reduction (ARR) is reported, these values are first converted to RR prior to generating $\alpha$ and $\beta$ paramaters.

$$SE_{CER} = \frac{\sqrt{CER \times (1 - CER)}}{n_c}$$

$$SE_{EER} = \frac{\sqrt{EER \times (1 - EER)}}{n_t}$$

$$\ln(RR) = \ln\left(\frac{EER}{CER}\right)$$

$$SE_{\ln(RR)} = \sqrt{\left(\frac{\text{SE}_{\text{EER}}}{\text{EER}}\right)^2 + \left(\frac{\text{SE}_{\text{CER}}}{\text{CER}}\right)^2}$$

$$CI_{RR} = e^{\ln(RR) \pm 1.96 \times SE_{\ln(RR)}}$$

$$RR = \frac{EER}{CER}$$

where:

- $EER$ is the Experimental Event Rate
- $CER$ is the Control Event Rate
- $n_c$ is the number of control/untreated patients
- $n_t$ is the number of treated patients

### 2 Models

#### 2.1 Model 1

Model 1 estimates the number of lives saved by CCP treatment compared to a scenario where CCP was not used. Total deaths were modeled as the sum of untreated and treated patients' deaths. Lives saved were calculated as the difference between the estimated deaths without CCP treatment and the observed deaths, using four mortality reduction scenarios (13%, 29%, 37%, and 47%) derived from both clinical trial and real-world data..

##### 2.1.1 Equations and Model Architecture

###### 2.1.1.1 Untreated Mortality Calculation

For each week ($i$) and each mortality reduction scenario ($j$), the untreated mortality rate ($U_m[i, j]$) is calculated based on the observed recorded deaths and the estimated reduction in mortality due to CCP:

$$U_m[i, j] = \frac{D[i]}{A[i] - T[i] \times M_r[j]} \tag{5}$$

where:

- $D[i]$ is the recorded deaths in week $i$, with lag.
- $A[i]$ is the total admissions in week $i$.
- $T[i]$ is the number of treated patients in week $i$.
- $M_r[j]$ is the mortality reduction scenario $j$.

###### 2.1.1.2 Mortality Reduction Prior

We specify prior distributions for the mortality reduction rates ($M_r$) using Beta distributions derived from Equation 3 and Equation 4:

$$M_r[j] \sim \text{Beta}(\alpha_j, \beta_j) \tag{6}$$

###### 2.1.1.3 Total Deaths Calculation

The total deaths ($TD[i,j]$) for each week ($i$) and each mortality reduction scenario ($j$) are calculated as:

$$TD[i,j] = (A[i] - T[i]) \times U_m[i,j] + T[i] \times U_m[i] \times (1 - M_r[j]) \tag{7}$$

###### 2.1.1.4 Likelihood

The likelihood of the observed recorded deaths ($D[i]$) given the estimated total deaths ($TD[i,j]$) is modeled as:

$$D[i] \sim \text{Normal}(TD[i,j], \sigma) \tag{8}$$

where $\sigma$ is the observed variability in weekly recorded deaths.

###### 2.1.1.5 Lives Saved Calculation

The number of lives saved by CCP treatment ($LS[i,j]$) each week is the difference between the total untreated deaths and the total deaths:

$$LS[i,j] = A[i] \times U_m[i,j] - TD[i,j] \tag{9}$$

###### 2.1.1.6 Summing Lives Saved

The estimated total lives saved by CCP treatment for each mortality reduction scenario is the sum of lives saved across all weeks:

$$LS_{total}[j] = \sum_{i=1}^{N} LS[i,j] \tag{10}$$

where $N$ is the number of weeks. The calculation is used across all models.

#### 2.2 Model 2

Model 2 estimates the number of lives saved by CCP treatment when administered within the first three days of hospitalization, reflecting optimal use. This model incorporates the reported usage rate from Mozzafari et al., which indicated that by December 2020, 83% of patients in a sample of US hospitals received CCP within the first three days of hospitalization. The model uses real-world efficacy data from Arnold et al., which reports a 47% reduction in mortality when CCP is given early.

##### 2.2.1 Equations and Model Architecture

###### 2.2.1.1 Untreated Mortality Calculation

For each week $(i)$, the untreated mortality rate $(U_m[i])$ is calculated based on the observed recorded deaths and the estimated reduction in mortality due to CCP:

$$U_m[i] = \frac{D[i]}{A[i] - T_e[i] \times M_r} \tag{11}$$

where:

- $D[i]$ is the recorded deaths in week $i$, with lag.
- $A[i]$ is the total admissions in week $i$.
- $T_e[i]$ is the number of early treated patients in week $i$.
- $M_r$ is the mortality reduction rate.

###### 2.2.1.2 Mortality Reduction Prior

Mortality reduction priors are defined as shown in Equation 6.

###### 2.2.1.3 CCP Usage Rate Prior

The prior for the CCP usage rate $(U_r)$ is also modeled using a Beta distribution based on the reported proportion of early CCP usage:

$$U_r \sim \text{Beta}(\alpha_{usage}, \beta_{usage}) \tag{12}$$

###### 2.2.1.4 Total Deaths Calculation

The total deaths $(TD[i])$ for each week $(i)$ are calculated as:

$$TD[i] = (A[i] - T_e[i]) \times U_m[i] + T_e[i] \times U_m[i] \times (1 - M_r) \tag{13}$$

###### 2.2.1.5 Likelihood

The likelihood of the observed recorded deaths ($D[i]$) given the total deaths ($TD[i]$) is modeled as a normal distribution with observed variability:

$$D[i] \sim \text{Normal}(TD[i], \sigma) \tag{14}$$

where $\sigma$ is the observed variability in weekly recorded deaths.

###### 2.2.1.6 Lives Saved

The number of lives saved by CCP treatment ($LS[i]$) each week is the difference between the total untreated deaths and the total deaths:

$$LS[i] = A[i] \times U_m[i] - TD[i] \tag{15}$$

#### 2.3 Model 3

Model 3 evaluates the hypothetical scenario in which all hospitalized patients received COVID convalescent plasma (CCP). This model assumes universal administration of CCP to estimate the number of lives saved, based on the four mortality reduction estimates used in Model 1. The total deaths in this model are calculated assuming all admissions received CCP, thereby estimating the treated mortality rate and total deaths accordingly.

##### 2.3.1 Equations and Model Architecture

###### 2.3.1.1 Untreated Mortality Calculation

Untreated mortality is defined the same as Equation 5.

###### 2.3.1.2 Treated Mortality Calculation

The treated mortality rate ($T_m[i, j]$) for each week ($i$) and each mortality reduction scenario ($j$) is calculated as:

$$T_m[i, j] = U_m[i, j] \times (1 - M_r[j])$$

###### 2.3.1.3 Likelihood

The likelihood of the observed recorded deaths ($D[i]$) is identitical to Equation 8.

###### 2.3.1.4 Estimated Deaths

The number of deaths if all patients were treated is estimated as:

$$D_t[i,j] = A[i] \times (1 - M_r[j])$$

###### 2.3.1.5 Lives Saved Calculation

The number of lives saved ($LS[i,j]$) each week is the difference between the number of recorded deaths and $D_t[i,j]$:

$$LS[i,j] = D[i] - D_t[i,j]$$

#### 2.4 Model 4

Model 4 is the same as model 3, but the calculation for the number of lives saved is:

$$LS[i,j] = A[i] \times U_m[i,j] - D_t[i,j]$$

#### 2.5 Model 5

Model 5 estimates the potential lives saved if COVID convalescent plasma (CCP) had been deployed for outpatient use. Given the greater efficacy of CCP when used early in the course of infection, it is likely that outpatient use could have saved more lives than inpatient use. The estimates are based on different reported efficacies in reducing hospitalization: 30%, 54%, and 80%. These figures were derived from various studies and meta-analyses conducted during the pandemic.

##### 2.5.1 Equations and Model Architecture

###### 2.5.1.1 Outpatient Efficacy Calculation

The efficacy of CCP in reducing hospitalization is modeled using a $Beta(\alpha, \beta)$ distribution. Three efficacy estimates are used: 30%, 54%, and 80%.

###### 2.5.1.2 Treated Mortality Rate Calculation

All cause mortality rate for hospitalized patients treated with CCP is treated as a *Beta* distribution with paramaters derived from Table 2 from Egloff et al. (J Clin Invest. 2021;131(20):e151788).

The mean was derived from the number of reported deaths out of the number of patients treated as a sample proportion.

From there,

$$Var = Mean \times \frac{1 - Mean}{n}$$

$$\alpha = \left( \frac{1 - \text{Mean}}{\text{Var}} - \frac{1}{\text{Mean}} \right) \times \text{Mean}^2$$

$$\beta = \alpha \times \left( \frac{1}{\text{Mean}} - 1 \right)$$

###### 2.5.1.3 Hospitalizations Avoided Calculation

The number of hospitalizations avoided ($HA[u, e]$) for each usage percentage ($u$) and each efficacy scenario ($e$) is calculated as:

$$HA[u, e] = \sum_{i=1}^{N} A[i] \times U[u] \times E[e]$$

where:

- $A[i]$ is the total admissions in week $i$.
- $U[u]$ is the usage percentage scenario $u$.
- $E[e]$ is the hospitalization reduction efficacy scenario $e$.

###### 2.5.1.4 Lives Saved Calculation

The number of lives saved ($LS[u, e]$) for each usage percentage ($u$) and each efficacy scenario ($e$) is calculated as:

$$LS[u, e] = \sum_{i=1}^{N} D[i] \times U[u] \times E[e]$$
